# Monitoring of the SARS-CoV-2 Omicron BA.1/BA.2 variant transition in the Swedish population reveals higher viral quantity in BA.2 cases

**DOI:** 10.1101/2022.03.26.22272984

**Authors:** Antonio Lentini, Antonio Pereira, Ola Winqvist, Björn Reinius

## Abstract

Throughout the SARS-CoV-2 pandemic, multiple waves of variants of concern have swept across populations, leading to a chain of new and yet more contagious lineages dominating COVID-19 cases. Here, we tracked the remarkably rapid shift from Omicron BA.1 to BA.2 sub-variant dominance in the Swedish population during January–March 2022. By analysis of 174,933 clinical nasopharyngeal swab samples using a custom variant-typing RT-PCR assay, we uncover nearly two-fold higher levels of viral RNA in cases with Omicron BA.2. Importantly, increased viral load in the upper pharynx upon BA.2 infection may provide part of the explanation why Omicron BA.2 is more transmissible and currently outcompetes the BA.1 variant across populations.

## Introduction

The SARS-CoV-2 pandemic has been characterised by the emergence and subsequent dominance of new variants of concern, such as Alpha B.1.1.7 during late 2020, Delta B.1.617.2 in mid 2021, and the Omicron variant B.1.1.529.1 (BA.1) in late 2021 and early 2022^1^. It is estimated that a peak of nearly 50 million new infections occurred daily world-wide during the Omicron wave of January 2022, far exceeding the peak of 14 million daily Delta infections during April 2021^2^ and signifying an unprecedented level of transmission of Omicron. In late December 2021, during the midst of the Omicron BA.1 wave in Sweden, the Omicron sub-variant BA.2 (B.1.1.529.2) arrived and rapidly spread through the population. At this time, we put in place an RT-PCR assay capable of genotyping Omicron BA.1 cases directly in primary SARS-CoV-2 RT-PCR testing, enabling day-by-day tracking of the BA.1 frequency at massive scale. Here, provide details of this assay and present data from 174,933 clinical nasopharyngeal swab samples analysed with the method in the Swedish population January–March 2022, demonstrating how the Omicron BA.1 wave was outcompeted by BA.2. Importantly, we further report approximately two-fold increased viral quantities detected in Omicron BA.2 cases, suggesting that higher viral load in the upper pharynx may at least partially explain why Omicron BA.2 is more contagious than the BA.1 lineage.

## Results and Discussion

We developed a modified version of the CDC SARS-CoV-2 RT-PCR assay^3^, simultaneously detecting general SARS-CoV-2 infection status (*N1-FAM*), human RNaseP sample integrity (*RP-HEX*), and Omicron BA.1-variant status (*S*_*BA1*_*-Cy5*) leveraging BA.1-specific indels in the spike (*S*) gene (**Fig. 1a**). To attain a multiplex assay of high sensitivity and specificity, suitable for RNA-extraction-free RT-PCR on heat-inactivated samples employed in mass testing^4^, we designed and evaluated 144 combinatorial *S*_*BA1*_*-*primer-probe sets, matching CDC *N1* properties and with minimized amplicon length (**Fig. S1a** and **Supplementary Table 1**). We obtained the first *in vitro* expanded Omicron BA.1 inoculate in Sweden (GISAID Accession ID: EPI_ISL_7452247) which was used as template, identifying Omicron BA.1-specific primer-probe sets (**Fig. 1a-b** and **Fig. S1b**). Detection sensitivity and log-linear cycle thresholds (C_T_) range were similar for the *N1* and *S*_*BA1*_ sets in the assay conditions (**Fig. 1c**). Using 185 clinical nasopharyngeal swab samples of known COVID19-infection status, we validated that addition of *S*_*BA1*_*-Cy5* probes did not affect *N1* C_T_ values and the sensitivity to detect general SARS-CoV-2 infection in primary RT-PCR (**Fig. 1d** and **Supplementary Table 2**). Parallel genotyping of 133 SARS-CoV-2-positive samples using Thermo Fisher TaqMan SARS-CoV-2 Mutation Panel Assay as well as whole-genome sequencing (WGS) provided Omicron-BA.1-case classification that was 100% consistent with our direct RT-PCR assay (**Fig1e-f**). Omicron (both the BA.1 and BA.2 lineage) carries a C>T substitution in the third base position of the *N1* probe, which we confirmed had negligible effect on C_T_ values by tests using CDC *N1* and custom Omicron *N1* probes (**Fig. S1c**).

**Figure 1.**
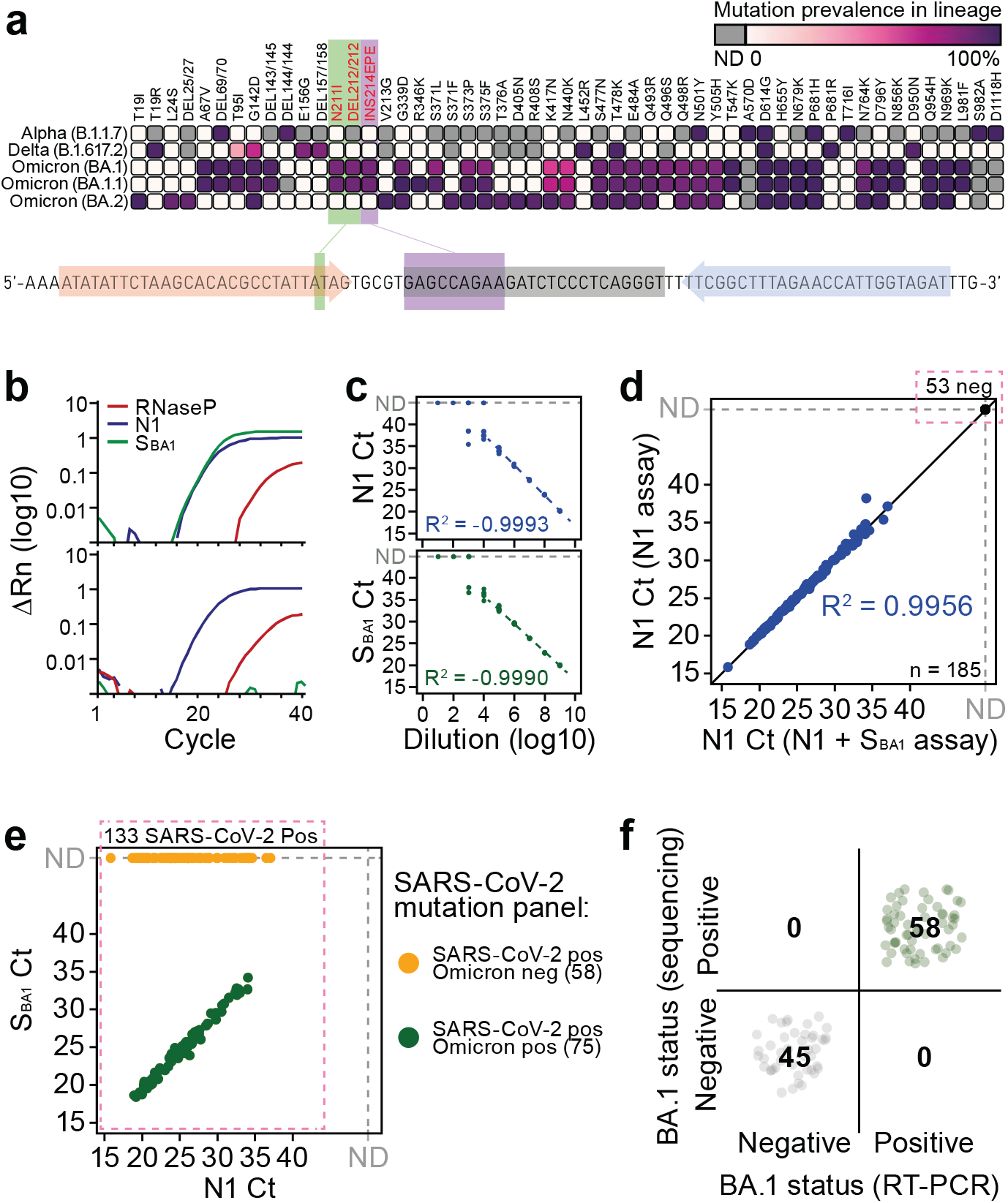
An RT-PCR assay providing general SARS-CoV-2 infection status and Omicron BA.1 classificaon in parallel. **a**. Mutaonal spectrum of the S protein among different SARS-CoV-2 lineages and location of Omicron BA.1-specific primer-probes used in the assay. **b**. RT-PCR amplification curves for N1, S_BA1_ and RNaseP targets in representave Omicron BA.1-positive (top) and BA.1 negative (boom) samples. **c**. RT-PCR sensivity of N1 (top) and S_BA1_ targets in dilution series of a BA.1-positive sample. Each dilution point represented by n = 8 replicates. **d**. Linear relationship between N1 RT-PCR Ct between the N1 assay and N1+S_BA1_ assay for n = 185 biological replicates. **e**. Agreement between lineage assignment of N1+S_BA1_ qPCR assay and Thermo Fisher TaqMan SARS-CoV-2 Mutation Panel (colours) for n = 133 COVID-19 positive samples (See also Supplementary Table 2). **f**. Agreement between N1+S_BA1_ RT-PCR assay and WGS lineage calls for n = 103 biological replicates.

Having optimised and validated the assay, it was deployed to monitor the Omicron BA.1 wave across central Sweden day-by-day, totalling 174,933 nasopharyngeal samples analysed between January 26 – March 8, 2022 (**Supplementary Table 3**). While BA.1 was the dominant variant among COVID19-positive cases during January, the BA.1 fraction steadily decreased to as low as 11% by March (**Fig. 2a**). This trend was observed in all monitored healthcare districts across central Sweden (**Fig. 2b**), indicating that Omicron BA.1 was being outcompeted by another lineage. To trace this variant switch, we whole-genome sequenced and assigned variants to 698 samples picked among SARS-CoV-2-positive cases. Strikingly, 100% of clinical samples typed as Omicron BA.1 negative in our direct RT-PCR assay were found to be of the Omicron BA.2 lineage by WGS (**Fig. 2c**), lacking the BA.1-specific “EPE” insertion in *S* gene targeted by our *S*_*BA1*_*-*probe (**Fig. S1a**), demonstrating that Omicron BA.2 was the variant outcompeting BA.1 in the Swedish population. Combining these clinically sequenced cases with the validation set of 133 samples mentioned in a previous section, we had classified a total of 801 cases by WGS (**Supplementary Table 4**), demonstrating >99% agreement with the BA.1 calls obtained directly in the RT-PCR (796/801 samples) (**Fig. 2d**). Why Omicron BA.2 is more contagious and outcompetes BA.1 is not well understood, indeed the BA.1/BA.2 transition is still ongoing across the world. Our massive RT-PCR dataset allowed us to compare viral levels detected in BA.1 and BA.2 cases. Intriguingly, samples genotyped as BA.1-negative in RT-PCR contained 1.9-fold higher levels of viral RNA than BA.1-positive samples (24.50 *vs*. 25.43 median *N1* C_T_; *P* = 1.54×10^−180^, Mann-Whitney U-test) (**Fig. 2e**), pointing to a substantial difference in viral load in the pharynx of patients infected with the two different Omicron variants. Importantly, this difference was observable across day-by-day timepoints (**Fig. S2c**) and was clearly attributable to the Omicron BA.2 variant, as BA.2 completely dominated non-BA.1 cases and viral copy-numbers were similar in Omicron BA.1 and Delta cases sampled in mid-December (*P* = 0.77, Mann-Whitney U-test) (**Fig. 2f**). Similar viral loads between Delta and BA.1 has previously been observed^5,6^ whereas the increased viral load in BA.2 cases was surprising. To ensure that the difference in viral quantity detected in samples of Omicron BA.1 and BA.2 cases was not attributed to our specific RT-PCR assay nor due to difference in amplification of the 3’-end of the SARS-CoV-2 genome (where the *N* gene is located) due to unequal discontinuous transcription found in coronaviruses^7^, we subjected 3,392 samples to both our *N1/RP/S*_*BA1*_ assay and an extraction-based assay (SARS-CoV-2 One-Step RT-PCR Kit, RdRp and N Genes, IVD, NZYTech; **Supplementary Table 5**) targeting *N* as well as *RdRp*, located further 5’ inside *ORF1ab*. Our RT-PCR assay for the *N* gene showed a strong linear correlation for both *N* and *RdRp* in the extraction-based assay (R^2^ > 0.949) (**Fig. S2d**) and we indeed confirmed increased copy numbers in BA.1-negative COVID19 samples in both assays and genes probed (**Fig. S2e**). Furthermore, lineage classification by WGS for 118 of these samples confirmed that BA.1-negative samples were indeed of BA.2 lineage in this analysis (81/81) (**Fig. S2f**).

**Figure 2.**
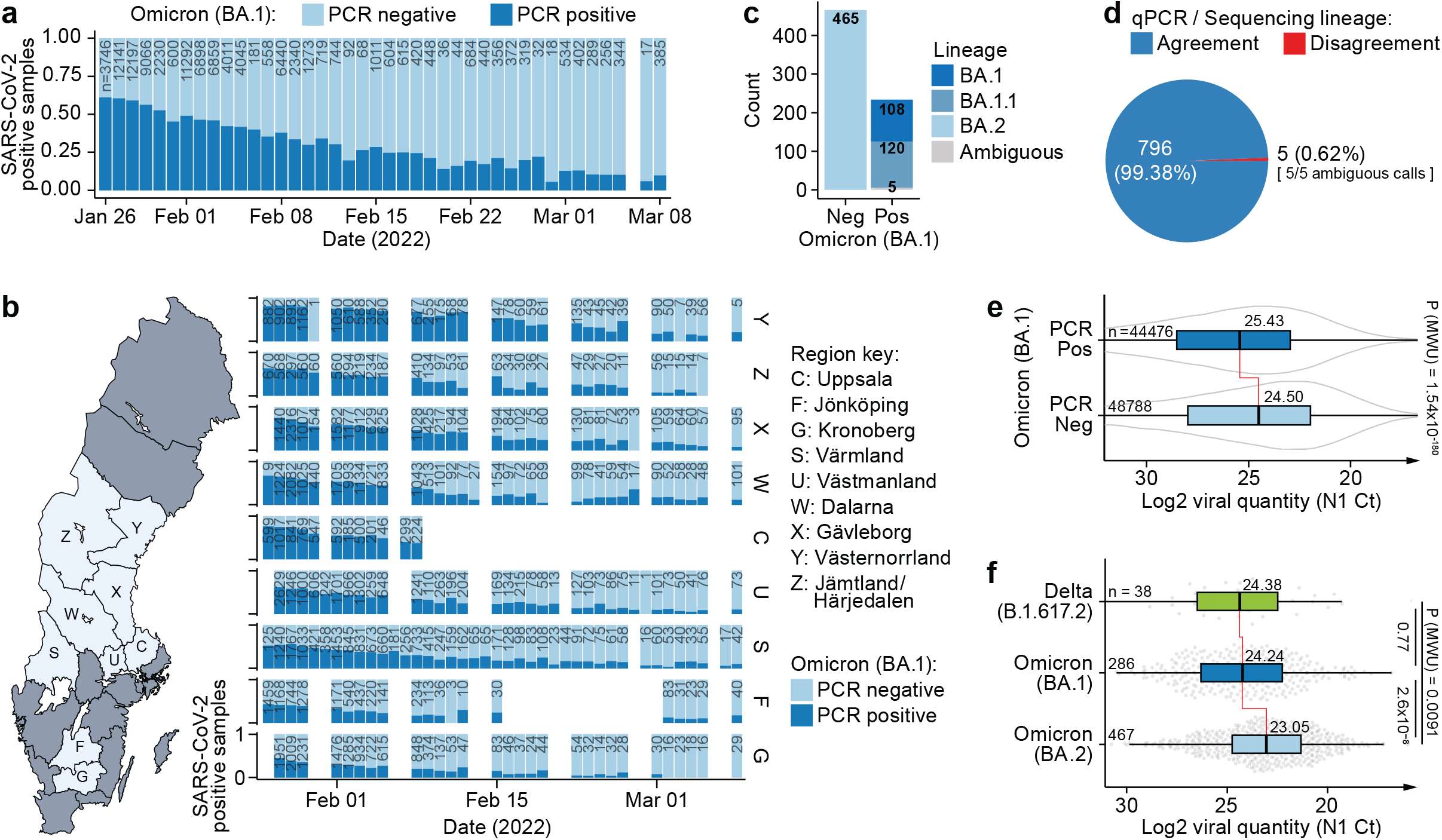
Omicron BA.1/BA.2 lineage transition and viral load in Swedish COVID-19 cases. **a**. Omicron BA.1 lineage assignment by RT-PCR over time in n = 93,126 SARS-CoV-2-positive cases (174,933 tests performed including negative cases). **b**. Same as **a**. but separated by originang Swedish healthcare districts. **c**. SARS-CoV-2 Lineage assignment by WGS based on qPCR lineage call (x-axis) for n = 698 biological replicates. **d**. Agreement between lineage calls for RT-PCR and WGS for n = 801 biological replicates. **e**. Difference in viral load (N1 RT-PCR Ct) for BA.1-positive and negative COVID-19 positive samples based on RT-PCR calls for n = 93,264 SARS-CoV-2-positive cases. P-values calculated using two-tailed Mann-Whitney U-tests. **f**. Same as **e**. but with classificaon based on WGS lineage calls for n = 791 biological replicates. P-values calculated using two-tailed Mann-Whitney U-tests.

Finally, we observed the BA.2-specific ORF3a:H78Y mutation (**Supplementary Table 6**), found primarily in Denmark^8^, in around 40% (172/425) of BA.2 cases classified by WGS (**Fig. S3a**). These cases were found almost exclusively in the southern-most Swedish region in our analysis where it accounted for around 72% of cases (103/144) (**Fig. S3b**), suggesting a common Danish/Swedish origin. We further found no Delta/Omicron recombination in our WGS data (**Fig. S3c**) which has been previously reported for few cases in southern Europe^9^.

Together, we present an effective RT-PCR assay to simultaneously and robustly call general COVID19 status and Omicron BA.1 variant status in a single RT-PCR reaction, which enabled real-time tracking of the Omicron BA.1/BA.2 transition in Sweden at immense scale. We observed that the predominant Omicron BA.1 variant was gradually and consistently outcompeted by Omicron BA.2 throughout Swedish healthcare districts. Importantly, we report that cases of BA.2 infection presented nearly two-fold higher quantities of viral RNA in nasopharyngeal sabs compared to BA.1 infection, suggesting that increased viral load in the upper pharynx may contribute to BA.2 being more contagious than the BA.1 lineage. Our direct Omicron BA.1-typing assay is a directly compatible addition to the well-established CDC *N1/RNaseP* probe sets and can thus easily be deployed for instant monitoring of the Omicron BA.1/BA.2 transition where needed, avoiding the time-lag and sample-number bottleneck of genotyping by WGS sequencing.

## Data Availability

All data produced in the present study are available upon reasonable request to the authors.

https://github.com/reiniuslab/OmicronWaves/

## Ethics statement

Ethical oversight and approval were obtained by the Swedish Ethical Review Authority (Dnr 2020-01945 and 2022-01139-02, Etikprövningsmyndigheten).

## Author contributions

AL performed WGS data processing, performed data analysis and visualisation, prepared figures, and wrote the manuscript. AP performed Thermo Fisher TaqMan SARS-CoV-2 Mutation Panel Assay. OW provided resources and coordinated the project. BR provided resources, coordinated the project, designed the BA.1 assay and performed RT-PCR, analysed the data, prepared figures, and wrote the manuscript. All authors approved the final version of the manuscript.

## Acknowledgements

We are grateful to members of the Reinius lab and ABC Labs, helping at various stages of the project. *In vitro* expanded Omicron BA.1 inoculate was kindly provided by the Public Health Agency of Sweden and we thank Sandra Söderholm and Shaman Muradrasoli facilitating transfer of this sample. We thank Jessica Alm at the National Pandemic Centre (NPC), Karolinska Institutet for providing anonymized control samples.

## Sequencing data and code availability

Raw sequencing data has been deposited to ArrayExpress (accession pending) and assembled SARS-CoV-2 genomes have been deposited to GISAID (accession pending). Computational code is available at https://github.com/reiniuslab/OmicronWaves/. All data are available upon request.

## Methods

### SARS-CoV-2 RT-PCR

The SARS-CoV-2 RT-PCR assay used in this study represents an improved, multiplex-version of our previously described RNA-extraction-free protocol^4^ with increased sample and reaction volume and increased sensitivity. For each reaction, 24 uL RT-PCR master mix was prepared, containing 7.5 μL TaqPath 1-Step RT-qPCR Master Mix, CG (Thermo Fisher, containing ROX as passive reference), 0.9 μL 10% Tween20 (Sigma), N1 primer-probe mix (IDT 2019-nCov CDC; FAM/BHQ1, Integrated DNA Technologies), S_BA1_ primer-probe mix (Cy-5/BHQ2; Merck), RNaseP primer-probe mix (HEX/BHQ1; Merck), and nuclease free water up to 24 uL. Primer/Probe concentrations in the final reactions were 246/62 nM (N1), 491/125 nM (S_BA1_), and 122/37 nM (RNaseP). Primer and probe sequences of the final assay are provided in **Supplementary Table 1b**. For RT-PCR testing, 6 μL heat-inactivated nasopharyngeal swab sample (in 0.9% saline) was added to optical 96-well PCR plates (EnduraPlate, Applied Biosystems) containing 24 μL master mix, using liquid a handling robot with automatic sample barcode scanner (Fluent 480, Fluent 780, or Fluent 1080; Tecan). Each PCR plate contained an Omicron BA.1 positive control sample and a negative control (water). Plates were sealed and centrifuged for 30 s at 1500 *g*. RT-PCR was performed on QuantStudio real-time PCR machines (Applied Biosystems) using the QuantStudio Design & Analysis Software v.1.5.2 and temperature cycles: 25 °C for 2 min, 50 °C for 10 min, 95 °C for 2 min, and 40 cycles of 95 °C for 3 s and 56 °C for 30 s. To test log-linear C_T_ range for the *N1* and *S*_*BA1*_ primer-probe sets in the assay conditions, a strongly positive Omicron BA.1 clinical sample was serial-diluted 1:10 in steps and RT-PCR was performed in 8 replicates per concentration. To test whether addition of the *S*_*BA1*_ primer-probe affected N1 C_T_s, we performed SARS-CoV-2 RT-PCR with and without presence of the S_BA1_ primer-probe set in parallel on 185 clinical samples (133 known positives, 52 negatives) and compared N1 C_T_s (**Fig. 1d**). In the parallel genotyping of 133 SARS-CoV-2-positive samples, performed using Thermo Fisher TaqMan SARS-CoV-2 Mutation Panel Assay (according to the manufacturers instructions) and our SARS-CoV-2 *N1/RP/S*_*BA1*_ assay, all Omicron samples were of the BA.1 lineage as demonstrated by WGS. Extraction-based SARS-CoV-2 One-Step RT-PCR Kit, RdRp and N Genes, IVD (NZYTech) was performed according to the manufacturer’s instructions (Version 13/2021, December 2021).

### SARS-CoV-2 samples

The in vitro expanded Omicron BA.1 reference sample used for the evaluation of S_BA1_ primer-probe sets (**Fig. S1a-b**) was obtained from the Public Health Agency of Sweden (sample isolated Nov 30, 2021; GISAID Accession ID: EPI_ISL_7452247), and the non-Omicron BA.1 reference sample (lacking “S:N211IN212-” and “S:214:EPE”) was an Alpha B.1.1.7 (sample isolated Feb 1, 2021; https://doi.org/10.5281/zenodo.4722502). 174,933 nasopharyngeal swab samples, collected in 0.9% saline solution and analysed January-March 2022 (**Supplementary Table 3**), were subjected to heat inactivation by 70 °C for 50 min in a hot air oven. These samples were analysed for presences of SARS-CoV-2 as part of clinical diagnostics performed at ABC Labs, Stockholm, by demand of the Public Health Agency of Sweden. The use of the Omicron BA.1 screening protocol was validated at ABC Labs and approved by the Public Health Agency of Sweden. For the purpose and analyses of the current study, sample identities were anonymised, and IDs were replaced by a randomized code (i.e., those listed in supplementary tables). Informed consent for the use of anonymized C_T_ values or samples obtained in routine clinical diagnostics was not obtained and is not required, and is in accordance with the study permit obtained by the Swedish Ethical Review Authority (Dnr 2020-01945 and 2022-01139-02, Etikprövningsmyndigheten).

### SARS-CoV-2 WGS

For sequencing of SARS-CoV-2 genomes, we used the Illumina COVIDSeq Test kit and sequencing was performed on the Illumina NextSeq 550 Sequencing System.

### Sequencing data analysis

Raw data was quality trimmed using fastp^10^ v0.20.0 and *de novo* assembled using MEGAHIT^11^ v1.2.9 [--min-contig-len 5000]. Disjoint contigs were scaffolded against the SARS-CoV-2 reference genome [ASM985889v3] using RagTag^12^ v2.1.0 and SARS-CoV-2 lineage was assigned using pangolin^13^ v3.1.20 [lineages version 2022-02-28]. Alignment statistics was obtained using Minimap2^14^ v2.24-r1122 [-ax sr Sars_cov_2.ASM985889v3.dna_sm.toplevel.fa.gz] and samtools^15^ v1.10 [stats] [coverage]. Identification of SARS-CoV-2 mutations was performed using Nextclade^16^v1.14.0. Computational code is available at https://github.com/reiniuslab/OmicronWaves/.

**Figure S1.**
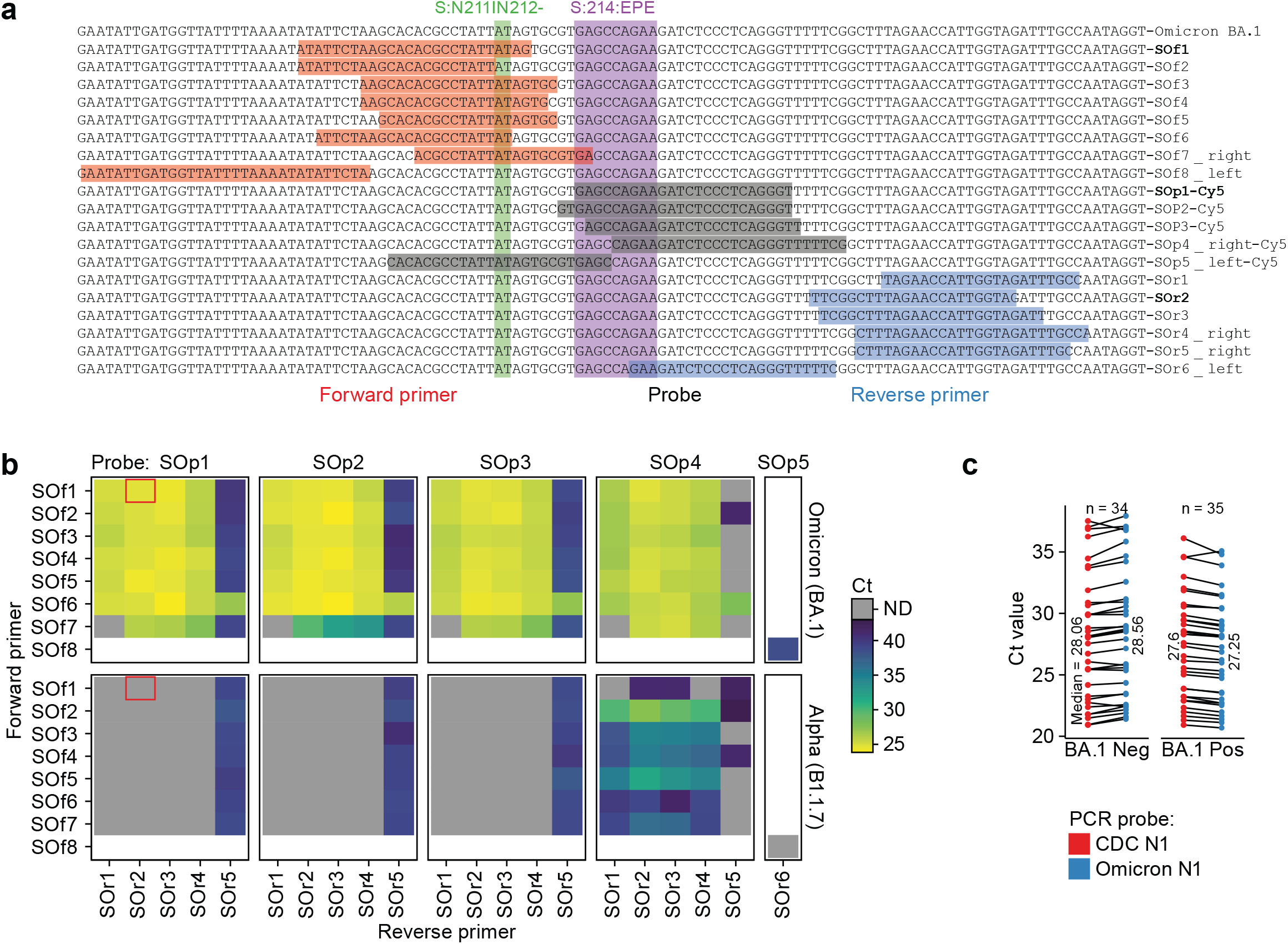
Extended analysis of Omicron BA.1-specific extraction-free RT-PCR. **a**. Evaluated primer-probe combinations targeng Omicron BA.1-specific mutations. **b**. Performance of primer-probe combinations in **a**. by RT-PCR. Red box indicates the selected primer-probe set. **c**. RT-PCR detection using the original CDC N1 probe or a custom Omicron-specific N1 probe designed around the C28311T mutation (present in both Omicron BA.1 and BA.2) located in the 3^rd^ base of the CDC N1 probe (See also Supplementary Table 1c). Data is stratified by BA.1 RT-PCR calls and shown as paired individual dots and median for n = 69 biological replicates.

**Figure S2.**
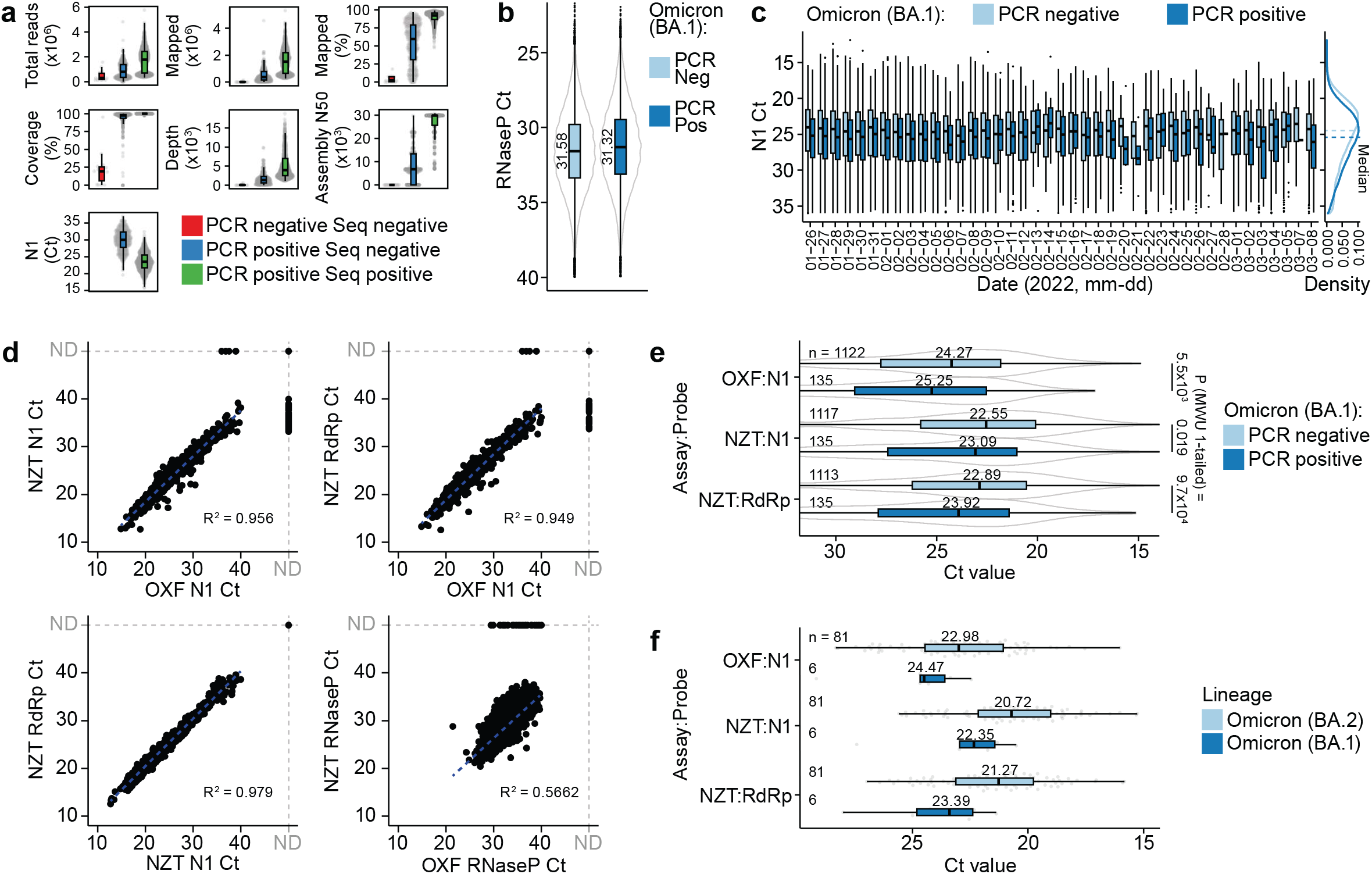
Extended analysis of WGS lineage calling results and viral load. **a**. Sequencing quality metrics separated by SARS-CoV-2 posivity by RT-PCR and successful lineage assignment by WGS for n = 1,153 biological replicates. **b**. RNaseP (human internal control) RT-PCR Ct values in samples stratified by BA.1 classification by RT-PCR for n = 93,264 SARS-CoV-2-positive cases (174,933 tests performed including negative cases). **c**. Viral load (N1 qPCR Ct) for BA.1-positive and negative SARS-CoV-2-positive samples based on RT-PCR calls over me for n = 93,126 biological replicates. Density plot with marked medians shown to the right. **d**. Linear relationship between Omicron extraction-free (N1+S_BA1_, OXF) and extraction-based (N1+RdRp, NZT) RT-PCR results for different primer-probe sets for n = 3,323 biological replicates. **e**. Difference in viral RNA quantity (RT-PCR Ct for different primer-probe sets) for BA.1-positive and negative SARS-CoV-2 positive samples based on RT-PCR calls for n = 1,252 biological replicates. OFX: Omicron extraction-free (N1+S_BA1_). NZT: Extraction-based SARS-CoV-2 One-Step RT-PCR Kit, RdRp and N Genes, IVD (NZYTech). P-values calculated using one-tailed Mann-Whitney U-tests. **f**. Same as **e**. but based on WGS lineage calls for n = 87 biological replicates.

**Figure S3.**
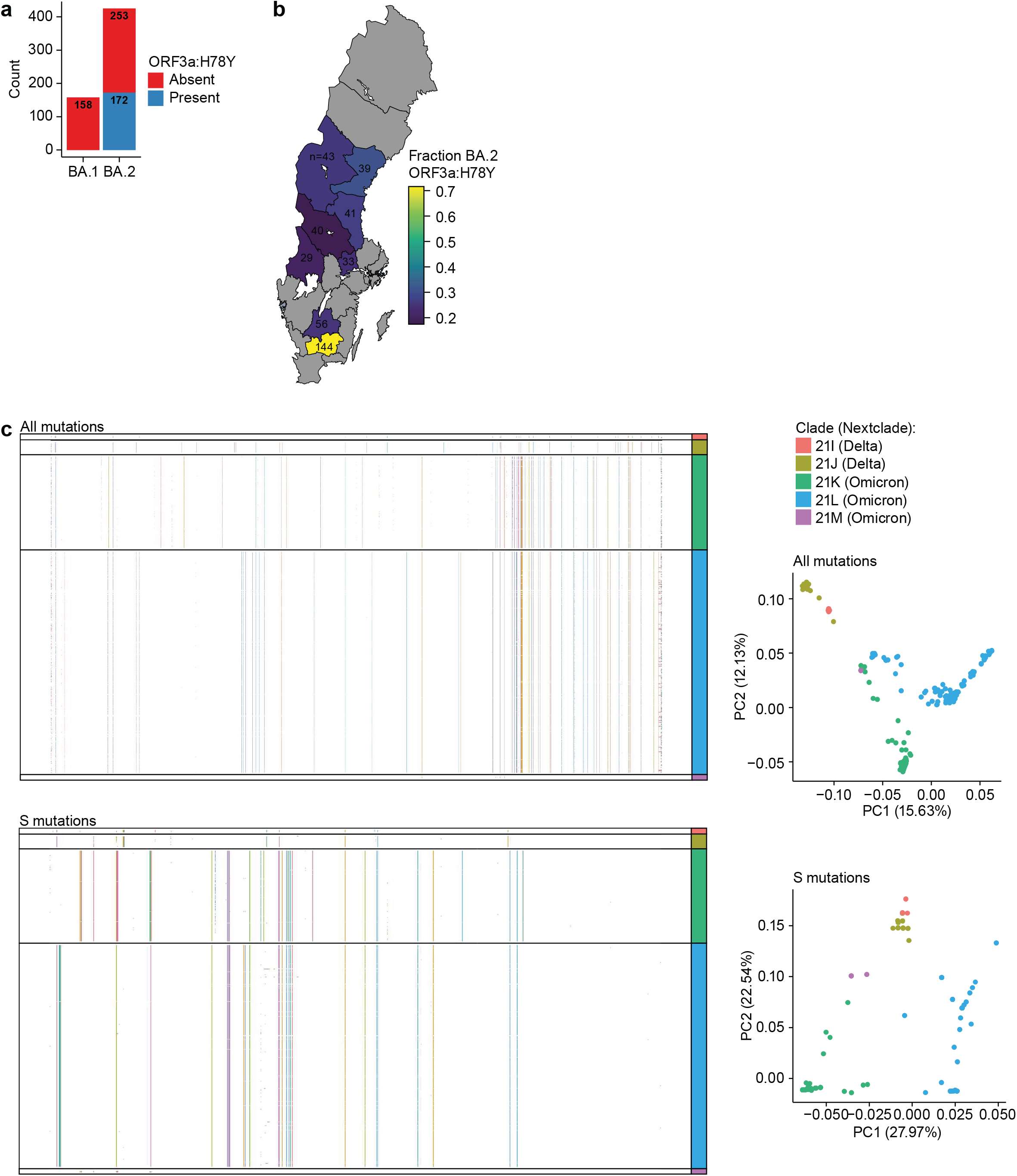
Extended analysis of SARS-CoV-2 mutations. **a**. Prevalence of ORF3a:H78Y mutations in BA.1- and BA.2 positive samples classified by WGS for n = 593 biological replicates. **b**. Same as **a**. but shown as mutation prevalence across Swedish healthcare regions. **c**. Full SARS-CoV-2 mutational spectrum (top) or S-specific mutations (bottom) for n = 712 biological replicates. Shown as heatmap with arbitrarily coloured mutations (left) or PCA analysis (right).

## References

1 World Health Organization. Tracking SARS-CoV-2 variants. https://www.who.int/en/activities/tracking-SARS-CoV-2-variants/ (accessed March 23, 2022).

2 Institute for Health Metrics and Evaluation. IHME | COVID-19 Projections. Institute for Health Metrics and Evaluation. https://covid19.healthdata.org/ (accessed March 23, 2022).

3 Lu X, Wang L, Sakthivel SK, et al. US CDC Real-Time Reverse Transcription PCR Panel for Detection of Severe Acute Respiratory Syndrome Coronavirus 2. Emerg Infect Dis 2020; 26. DOI:10.3201/eid2608.201246.

4 Smyrlaki I, Ekman M, Lentini A, et al. Massive and rapid COVID-19 testing is feasible by extraction-free SARS-CoV-2 RT-PCR. Nat Commun 2020; 11: 4812.

5 Hay JA, Kissler SM, Fauver JR, et al. Viral dynamics and duration of PCR positivity of the SARS-CoV-2 Omicron variant. Epidemiology, 2022 DOI:10.1101/2022.01.13.22269257.

6 Puhach O, Adea K, Hulo N, et al. Infectious viral load in unvaccinated and vaccinated patients infected with SARS-CoV-2 WT, Delta and Omicron. Infectious Diseases (except HIV/AIDS), 2022 DOI:10.1101/2022.01.10.22269010.

7 Sawicki SG, Sawicki DL. Coronaviruses use Discontinuous Extension for Synthesis of Subgenome-Length Negative Strands. In: Talbot PJ, Levy GA, eds. Corona-and Related Viruses. Boston, MA: Springer US, 1995: 499–506.

8 Desingu PA, Nagarajan K. Omicron BA.2 lineage spreads in clusters and is concentrated in Denmark. Journal of Medical Virology 2022; : jmv.27659.

9 Colson P, Fournier P-E, Delerce J, et al. Culture and identification of a “Deltamicron” SARS-CoV-2 in a three cases cluster in southern France. Infectious Diseases (except HIV/AIDS), 2022 DOI:10.1101/2022.03.03.22271812.

10 Chen S, Zhou Y, Chen Y, Gu J. fastp: an ultra-fast all-in-one FASTQ preprocessor. Bioinformatics 2018; 34: i884–90.

11 Li D, Liu C-M, Luo R, Sadakane K, Lam T-W. MEGAHIT: an ultra-fast single-node solution for large and complex metagenomics assembly via succinct de Bruijn graph. Bioinformatics 2015; 31: 1674–6.

12 Alonge M, Lebeigle L, Kirsche M, et al. Automated assembly scaffolding elevates a new tomato system for high-throughput genome editing. Plant Biology, 2021 DOI:10.1101/2021.11.18.469135.

13 O’Toole Á, Scher E, Underwood A, et al. Assignment of Epidemiological Lineages in an Emerging Pandemic Using the Pangolin Tool. Virus Evolution 2021; : veab064.

14 Li H. Minimap2: pairwise alignment for nucleotide sequences. Bioinformatics 2018; 34: 3094–100.

15 Li H, Handsaker B, Wysoker A, et al. The Sequence Alignment/Map format and SAMtools. Bioinformatics 2009; 25: 2078–9.

16 Aksamentov I, Roemer C, Hodcroft E, Neher R. Nextclade: clade assignment, mutation calling and quality control for viral genomes. JOSS 2021; 6: 3773.

